# Immediate recruitment of dormant coronary collaterals can provide more than half of normal resting perfusion during coronary occlusion in patients with coronary artery disease

**DOI:** 10.1101/2021.10.07.21264445

**Authors:** Brandon J Reid, Thomas Lindow, Stafford Warren, Eva Persson, Ravinay Bhindi, Michael Ringborn, Martin Ugander, Usaid Allahwala

**Affiliations:** Kolling Institute, Department of Cardiology, Royal North Shore Hospital, and University of Sydney, Sydney, Australia; Clinical Sciences, Department of Clinical Physiology, Skane University Hospital, Lund, Sweden; Anne Arundel Medical Center, Department of Medicine, Cardiology Division, Annapolis, MD, USA; Thoracic Center, Blekinge County Hospital, Karlskrona, Sweden

**Keywords:** Coronary angiography, coronary physiology, collateral flow

## Abstract

**Aims:** Dormant coronary collaterals are highly prevalent and clinically beneficial in cases of coronary occlusion. However, the magnitude of myocardial perfusion provided by immediate coronary collateral recruitment during acute occlusion is unknown. The aim of this study was to quantify collateral myocardial perfusion during balloon occlusion in patients with coronary artery disease (CAD).

**Methods and results:** Patients without angiographically visible collaterals undergoing elective percutaneous transluminal coronary angioplasty (PTCA) to a single epicardial vessel underwent two scans with ^99m^Tc-sestamibi myocardial perfusion single-photon emission computed tomography (SPECT). All subjects underwent at least three minutes of angiographically verified complete balloon occlusion, at which time an intravenous injection of the radiotracer was administered, followed by SPECT imaging. A second radiotracer injection followed by SPECT imaging was performed 24 hours after PTCA.

The study included 22 patients (median [interquartile range] age 68 [54-72] years, 10 (45%) female). The perfusion defect extent was 19 [11–38] % of the LV, and the collateral perfusion at rest was 64 [58-67]% of normal.

**Conclusion:** This is the first study to describe the magnitude of short-term changes in coronary microvascular collateral perfusion in patients with CAD. On average, despite coronary occlusion and an absence of angiographically visible collateral vessels, collaterals provided more than half of the normal perfusion.

**Graphical abstract:** 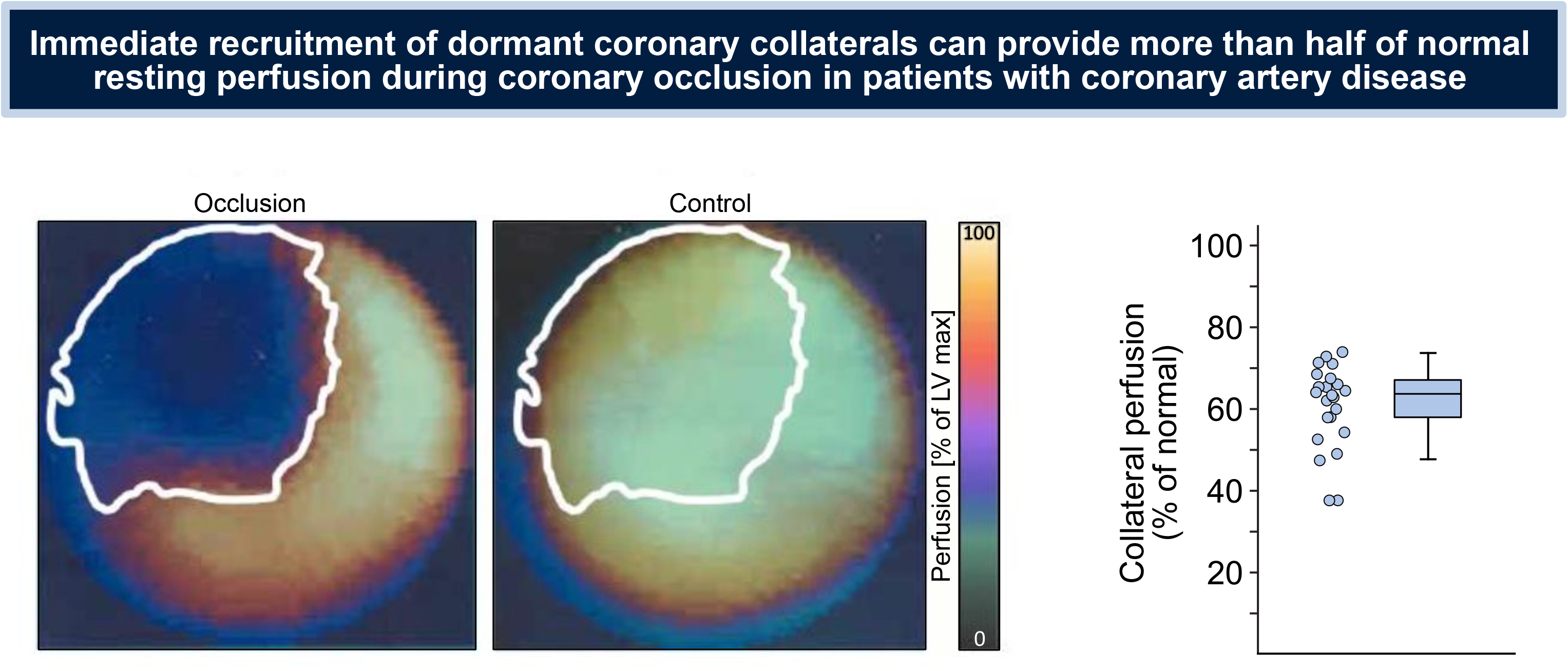

Dormant collaterals are highly prevalent but the magnitude of myocardial perfusion provided by immediate coronary collateral recruitment during acute occlusion in humans is unknown. Patients without angiographically visible collaterals underwent intravenous injection with 99mTc-sestamibi during coronary balloon inflation followed by SPECT imaging. A control scan was performed 24 hours later..Median collateral perfusion during coronary occlusion was 64% of normal.

## Introduction

The coronary collateral circulation is a preformed network of anastomotic connections between primitive vessels, linking one epicardial artery with another, acting as a “natural bypass” mechanism [1, 2]. The presence of robust coronary collateral circulation is known to be associated with improved survival and left ventricular function in the setting of ST-elevation myocardial infarction (STEMI) [3].

Whilst the prevalence of collaterals varies between species, approximately 25% of patients have angiographically visible robust collaterals at the time of ST elevation myocardial infarction [4]. Short-term changes in collateral flow in humans has been described angiographically by Rentrop et al [1]. In that study, patients with stable angina and single-vessel disease were studied by contralateral coronary artery contrast injection during balloon inflation. Most patients had no collateral filling at baseline, while all but one patient showed improved collateral filling upon balloon occlusion. This suggests a high prevalence of dormant collaterals, but does not provide reliable information on magnitude of the resultant perfusion.

Importantly, collateral vessels smaller than approximately 100 μm are not visibly detectable by invasive angiography [5]. However, relative myocardial perfusion can be quantified by ^99m^Tc-sestamibi single-photon emission computed tomography (SPECT) regardless of vessel size. ^99m^Tc-sestamibi uptake is proportional to myocardial perfusion [6], and it exhibits minimal redistribution after initial uptake [7]. While the contribution of collateral flow has been thoroughly described in canine models using microsphere techniques [8-13], there is no quantitative data on short-term changes in collateral perfusion in man. Of note, infarct biology cannot be readily translated from canine models to man [14], and an understanding of human basic coronary physiology is essential to cardiovascular medicine. Therefore, the aim of the study was to quantify collateral myocardial perfusion during experimental coronary balloon occlusion in patients with CAD using ^99m^Tc-sestamibi SPECT.

## Methods

### Study design

The current study was a retrospective substudy of a previously published larger cohort [15]. In that study, 42 patients with obstructive, single-vessel coronary artery disease underwent elective prolonged balloon inflation during percutaneous transluminal coronary angioplasty (PTCA), and two myocardial perfusion single-photon emission computed tomography (SPECT) scans between September 1995 and April 1996 with radioactive tracer injections during coronary occlusion and at rest the subsequent day [15]. Due to either technical problems, unsuccessful injections, or prior coronary artery bypass surgery to the balloon-occluded artery, 7 patients were excluded from the original study. For the purpose of this study, patients with angiographically visible collaterals, prior infarction, or coronary artery bypass surgery (not only to the balloon-occluded artery) were further excluded (n=13). The study was undertaken at Charleston Area Medical Center, Charleston, West Virginia, USA, and was approved by the local Investigational Review Board. All subjects provided written informed consent.

### SPECT Imaging

For each patient, when the angioplasty balloon was angiographically verified as being fully inflated during PTCA, with absence of anterograde blood flow, 1100 MBq (30 mCi) ^99m^Tc-sestamibi was injected intravenously. The intracoronary balloon inflation lasted for at least three minutes in all patients, in order to maximize clearance of ^99m^Tc-sestamibi from the bloodstream.

Immediately following PTCA, the interventional cardiologist performing the procedure recorded the location of balloon occlusion and the duration of balloon inflation. Also, at the conclusion of the procedure, the interventional cardiologist noted whether angiographically visible collateral circulation was considered to be present upon review of the angiographic images of all coronary vessels. Within three hours following angioplasty, the patient was brought to the nuclear medicine laboratory to acquire SPECT imaging data (Occlusion study). On the day following PTCA, a second intravenous injection with 1100 MBq of ^99m^Tc-sestamibi was administered intravenously, followed by SPECT imaging within three hours (Control study). Each patient in the study was clinically stable throughout both processes.

SPECT imaging was performed using a single-head gamma camera (Elscint, Haifa, Israel) with a 140 keV (±20%) energy window. Images were acquired using a low energy high-resolution collimator in a 64×64 matrix, 6.9 mm pixel size, using 30 projections (25 seconds/projection) over 180° from 45° right anterior oblique to 45° left posterior oblique. A filtered back projection with a Butterworth filter (order 5, cut-off 0.25 cycles/pixel) was used to reconstruct axial slices. The same gamma camera and image acquisition procedures were used for the Occlusion and Control studies.

### Image Analysis

Short-axis images were reconstructed using the Cedars-Emory quantitative analysis software (CEqual), and were used to build a volume-weighted bull’s eye plots [16]. The quantification of collateral perfusion was performed by comparing the Occlusion study to the Control study for each patient. The Control study was first corrected by taking the decay of ^99m^Tc into account (24 h, Tc^99m^ decay constant λ= 0.1151 h^-1^). Following this process, the Occlusion study was scaled so that its average value, in the region above 90% of its maximum, was made equal to the average value of the control study in the same region of the heart. This normalization was necessary to achieve uniformity for the Control and Occlusion studies in regions not impacted by inflation-induced hypoperfusion.

The amount of residual perfusion downstream of the balloon-occluded artery, presumably due to coronary collateral flow, was quantified within the region of hypoperfusion in the following manner. First, the ratio of perfusion between the occlusion study and the control study was calculated. Secondly, the extent of the perfusion defect was identified in an extent map, and the region where the ratio of Occlusion-to-Control counts was less than 75% of tracer uptake was delineated, as previously validated [15]. Residual collateral perfusion, i.e. the perfusion within the perfusion defect in % of the normal perfusion within that same region, was calculated as Occlusion counts divided by Control counts (%). A representative case is illustrated in Figure 1.

**Figure 1.** Visualization of collateral perfusion to myocardium during elective PTCA in patients with CAD. Bull’s eye plots show left ventricular (LV) myocardial perfusion by ^99m^Tc-sestamibi SPECT in a representative patient with a balloon-occluded left anterior descending artery (Occlusion) and 24 hours later at rest (Control). The size of the perfusion defect (white delineation) was defined as the region where the Occlusion/Control ratio was less than 75% as previously validated [4]. Collateral perfusion within the perfusion defect was quantified as Occlusion counts divided by Control counts, expressed in percent.

### Statistical Analysis

Non-normally distributed data are presented as median [interquartile range]. Differences between groups were tested using the Wilcoxon test. Linear correlations were described using Spearman’s rank correlation coefficient <u>ρ</u>. A two-sided p-value less than 0.05 was considered statistically significant. All statistical analysis was performed using the software R (version 4.0.4, R Core Team, R Foundation for Statistical Computing, Vienna, Austria).

## Results

The study included 22 patients (age 68 [54-72] years, 10 (45%) female). The degree of diameter stenosis of treated vessels ranged from 60-99%, with successful PTCA performed with a mean balloon occlusion time of 5 minutes (range 3-7 minutes), resulting in ≤20% residual stenosis in all cases. Ten patients underwent stent implantation as part of the procedure. The vessels undergoing PTCA were 6 (27%) in the left anterior descending artery (LAD) [4 proximal], 7 (32%) in the left circumflex coronary artery (LCx) [5 proximal] and 9 (41%) in the right coronary artery (RCA) [4 proximal].

For the entire cohort, the median extent of the perfusion defect was 19 [11–38] % of the left ventricle. The extent of perfusion defects was larger in patients with LAD occlusion (47 [42–50] %), compared to RCA or LCx occlusion (14 [7–22] %), p<0.001. The residual collateral perfusion was 64 [58–67] % [Figure 2], 51 [48–54]% in LAD occlusion, and 65 [63–69]% in RCA or LCx occlusion (p<0.001). Collateral perfusion was negatively correlated with perfusion defect size (<u>ρ</u>=-0.82, p<0.001), but did not differ by sex (males 62 [54–66] vs females 65 [61– 68] %, p=0.34) and was not correlated to age (<u>ρ</u>=-0.18, p=0.42).

**Figure 2.** Quantification of collateral perfusion to myocardium during elective PTCA in patients with CAD. Box and whiskers plot show the collateral perfusion in all patients (n=22, median [interquartile range], 64 [58-67] %).

## Discussion

The main finding of the study was that immediate recruitment of dormant collaterals after balloon occlusion of an epicardial coronary artery results in substantial myocardial perfusion, averaging 64% of normal perfusion. This is the first study to describe the magnitude of short-term changes in collateral perfusion in human subjects with CAD. A high prevalence of short-term changes in collateral flow was described angiographically by Rentrop, et al, using contralateral contrast injections during balloon inflation, with improvement in angiographic collateral filling grade in all but one patient [1]. We add to this knowledge by providing quantification of the resulting myocardial perfusion. Our findings are in agreement with previous findings that collateral flow, in some cases, can be enough to prevent short-term ischemia and anginal symptoms [17].

### Collateral coronary perfusion in humans

The magnitude of collateral perfusion in our study was greater than previously described. Collateral perfusion has been evaluated in patients with successfully reperfused STEMI, and found to provide incremental information on final infarct size, beyond myocardium at risk, time to reperfusion, and infarct location [18]. In that study, 89 patients were injected with ^99m^Tc-sestamibi prior to revascularization by PTCA or thrombolysis. Collateral myocardial perfusion was estimated by comparing the counts within the perfusion defect to the maximal value within the same short-axis slice, and was found to be approximately 27%, which is less than in our study. Of note, in contrast to our methodology, the authors did not evaluate the collateral perfusion in reference to the normal perfusion within the affected myocardial region, but rather it was compared to remote myocardium in the same slice. Furthermore, that study reported the magnitude of collateral perfusion in a clinical cohort before emergent revascularization, after on average 5 hours of acute coronary occlusion and ongoing myocardial infarction. In contrast to the results of that study, the current is the first study to report the magnitude of perfusion in relation to normal perfusion that can be accomplished by collateral vessel recruitment in an immediate (3-5 minutes) response to coronary occlusion without the cofounding pathophysiology of relatively longstanding ongoing acute myocardial infarction.

### Collateral perfusion in animal models

By comparison, Reimer, et al, have reported from experimental studies in the dog, that collateral flow decreases over time in coronary occlusion, especially in subendocardial regions [8, 12]. The magnitude of collateral perfusion in humans, as shown in our study, is higher than in animal models [8-10]. Reimer, et al, found variable but consistently lower collateral flow in dogs with a proximally occluded LCx, ranging from 12 to 31% in subendocardial vs. subepicardial layers of the myocardium, in reference to normally perfused myocardium [8], and even values as low as 6% have been reported [10]. By comparison, the current study found more than twice greater collateral perfusion in patients with CAD, likely due to a combination of biological differences between species, age, and the presence of CAD.

### Technical considerations

The current study showed a negative correlation between perfusion defect size and the magnitude of collateral perfusion. In other words, smaller perfusion defects had greater collateral perfusion than larger perfusion defects. Phantom studies have shown that residual activity in perfusion defects is influenced not only by the collateral flow to that region, but also by pixel size, collimator resolution, type of orbit, defect size, and location. With the SPECT technology used in this study, a significant overestimation can be expected in defect sizes <10%, but the effect is of less importance with larger defect sizes [19]. By excluding those with an extent <10% the median residual collateral perfusion was still substantial (62%). The difference in the extent of perfusion defects between LAD and RCA or LCx occlusions likely contributes to the observed differences in residual perfusion between different ischemic locations. Taken together, these results suggest that although technical aspects can have influenced the results, there may be a limit to the distance over which collaterals can effectively perfuse into the core of a perfusion defect.

### The clinical importance of collaterals

Having robust coronary collateral circulation as visualized by invasive angiography has been associated with clinical benefits, since robust collaterals allow oxygenated blood to reach the jeopardized myocardium [2, 3, 20]. Patients with robust collaterals presenting with STEMI have a lower mortality in both the short and long term, and higher left ventricular ejection fraction [3]. Furthermore, patients with robust collaterals are more likely to have successful percutaneous coronary intervention to treat chronic total occlusion [20]. Future studies on coronary collateral circulation could incorporate quantification of the myocardial perfusion contributed by collaterals using perfusion imaging methods such those used in the current study.

### Limitations

The data used in this study stem from a study performed during the years 1995-1996, and technical SPECT advances have been made since then, which may affect the results compared to if the study had been undertaken with today’s SPECT technology. In addition, the sample included in this study is small. However, the access to data from complete epicardial coronary artery balloon occlusion and simultaneous radioactive tracer injection with control imaging in human subjects is unique and unlikely to be replicated. If the results are cautiously interpreted, significant knowledge can be gained from these records, despite apparent limitations.

Uptake of ^99m^Tc-sestamibi following balloon deflation could potentially falsely increase the estimation of collateral perfusion. However, <5% of the activity remains in the blood stream at 5 minutes after intravenous injection [21]. Since mean balloon injection time in our study was 5 minutes, this likely had minimal impact on our results.

Furthermore, contralateral vessel angiography was not performed during balloon inflation in this study. Thus, it is only known that collateral vessels could not be visualized prior to balloon occlusion, but it is not known if collateral vessels could be angiographically visualized during balloon occlusion, which otherwise would be the case for example in the assessment of chronic total coronary occlusion. Regardless, the angiographic visualization of collateral vessels was performed under clinical conditions, and the current results highlight that angiographic visualization under clinical conditions does not visualize a sizable portion of perfusion that can reach myocardium subtended by a given artery should it become occluded. Also, SPECT images were not corrected for attenuation, which may affect accuracy, albeit not substantially [22], and with minimal impact since the same attenuating properties would be at play during both the Occlusion and Control study.

## Conclusions

This is the first study to quantify the myocardial perfusion after immediate collateral recruitment following coronary occlusion in patients with CAD. On average, collaterals provided more than half of normal myocardial perfusion in patients with CAD. Future research on diagnosis and therapy in CAD could consider quantification of collateral perfusion beyond angiographic visualization.

## Data Availability

Data can be made available upon reasonable request to the corresponding author.

## Acknowledgements

The authors thank medical physicist Dr. Enid Eslick, PhD, for valuable discussion and input regarding nuclear medicine physics-related aspects of the methodology.

## References

1. Rentrop KP, Cohen M, Blanke H, et al. Changes in collateral channel filling immediately after controlled coronary artery occlusion by an angioplasty balloon in human subjects. J Am Coll Cardiol. 1985;5:587–92.

2. Rentrop KP, Feit F, Sherman W, et al. Serial angiographic assessment of coronary artery obstruction and collateral flow in acute myocardial infarction. Report from the second Mount Sinai-New York University Reperfusion Trial. Circulation. 1989;80:1166–75.

3. Allahwala UK, Nour D, Alsanjari O, et al. Prognostic implications of the rapid recruitment of coronary collaterals during ST elevation myocardial infarction (STEMI): a meta-analysis of over 14,000 patients. Journal of thrombosis and thrombolysis. 2021;51:1005–16.

4. Allahwala UK, Weaver JC, Nelson GI, et al. Effect of Recruitment of Acute Coronary Collaterals on In-Hospital Mortality and on Left Ventricular Function in Patients Presenting With ST Elevation Myocardial Infarction. Am J Cardiol. 2020;125:1455–60.

5. Gensini GG,Bruto da Costa BC. The coronary collateral circulation in living man. Am J Cardiol. 1969;24:393–400.

6. Sinusas AJ, Trautman KA, Bergin JD, et al. Quantification of area at risk during coronary occlusion and degree of myocardial salvage after reperfusion with technetium-99m methoxyisobutyl isonitrile. Circulation. 1990;82:1424–37.

7. Haronian HL, Remetz MS, Sinusas AJ, et al. Myocardial risk area defined by technetium-99m sestamibi imaging during percutaneous transluminal coronary angioplasty: comparison with coronary angiography. J Am Coll Cardiol. 1993;22:1033–43.

8. Reimer KA, Jennings RB, Cobb FR, et al. Animal models for protecting ischemic myocardium: results of the NHLBI Cooperative Study. Comparison of unconscious and conscious dog models. Circulation research. 1985;56:651–65.

9. Murdock RH, Jr., Chu A, Grubb M, et al. Effects of reestablishing blood flow on extent of myocardial infarction in conscious dogs. Am J Physiol. 1985;249:H783–91.

10. Ugander M, Bagi PS, Oki AJ, et al. Myocardial edema as detected by pre-contrast T1 and T2 CMR delineates area at risk associated with acute myocardial infarction. JACC Cardiovasc Imaging. 2012;5:596–603.

11. Reimer KA,Jennings RB. The “wavefront phenomenon” of myocardial ischemic cell death. II. Transmural progression of necrosis within the framework of ischemic bed size (myocardium at risk) and collateral flow. Laboratory investigation; a journal of technical methods and pathology. 1979;40:633–44.

12. Reimer KA, Lowe JE, Rasmussen MM, et al. The wavefront phenomenon of ischemic cell death. 1. Myocardial infarct size vs duration of coronary occlusion in dogs. Circulation. 1977;56:786–94.

13. Jennings RB, Ganote CE,Reimer KA. Ischemic tissue injury. Am J Pathol. 1975;81:179–98.

14. Hedström E, Engblom H, Frogner F, et al. Infarct evolution in man studied in patients with first-time coronary occlusion in comparison to different species - implications for assessment of myocardial salvage. Journal of cardiovascular magnetic resonance : official journal of the Society for Cardiovascular Magnetic Resonance. 2009;11:38-.

15. Persson E, Palmer J, Pettersson J, et al. Quantification of myocardial hypoperfusion with 99m Tc-sestamibi in patients undergoing prolonged coronary artery balloon occlusion. Nucl Med Commun. 2002;23:219–28.

16. Garcia EV, Cooke CD, Van Train KF, et al. Technical aspects of myocardial SPECT imaging with technetium-99m sestamibi. Am J Cardiol. 1990;66:23e–31e.

17. Wustmann K, Zbinden S, Windecker S, et al. Is there functional collateral flow during vascular occlusion in angiographically normal coronary arteries? Circulation. 2003;107:2213–20.

18. Christian TF, Gibbons RJ, Clements IP, et al. Estimates of myocardium at risk and collateral flow in acute myocardial infarction using electrocardiographic indexes with comparison to radionuclide and angiographic measures. J Am Coll Cardiol. 1995;26:388–93.

19. O’Connor MK, Gibbons RJ, Juni JE, et al. Quantitative myocardial SPECT for infarct sizing: feasibility of a multicenter trial evaluated using a cardiac phantom. Journal of nuclear medicine : official publication, Society of Nuclear Medicine. 1995;36:1130–6.

20. Allahwala UK, Nour D, Bhatia K, et al. Prognostic impact of collaterals in patients with a coronary chronic total occlusion: A meta-analysis of over 3,000 patients. Catheter Cardiovasc Interv. 2021;97:E771–e7.

21. Boschi A, Uccelli L, Marvelli L, et al. Technetium-99m Radiopharmaceuticals for Ideal Myocardial Perfusion Imaging: Lost and Found Opportunities. Molecules. 2022;27.

22. Slomka PJ, Fish MB, Lorenzo S, et al. Simplified normal limits and automated quantitative assessment for attenuation-corrected myocardial perfusion SPECT. J Nucl Cardiol. 2006;13:642–51.

